# Potential impact of outpatient stewardship interventions on antibiotic exposures of bacterial pathogens

**DOI:** 10.1101/19008029

**Authors:** Christine Tedijanto, Yonatan H Grad, Marc Lipsitch

**Affiliations:** Center for Communicable Disease Dynamics, Department of Epidemiology, Harvard T.H. Chan School of Public Health, Boston, Massachusetts, USA; Department of Immunology and Infectious Diseases, Harvard T.H. Chan School of Public Health, Boston, Massachusetts, USA; Division of Infectious Diseases, Brigham and Women’s Hospital, Harvard Medical School, Boston, Massachusetts, USA

## Abstract

The relationship between antibiotic stewardship and population levels of antibiotic resistance remains unclear. In order to better understand shifts in selective pressure due to stewardship, we use publicly available data to estimate the effect of changes in prescribing on exposures to frequently used antibiotics experienced by potentially pathogenic bacteria that are asymptomatically colonizing the microbiome. We quantify this impact under four hypothetical stewardship strategies. In one scenario, we estimate that elimination of all unnecessary outpatient antibiotic use could avert 6 to 48% (IQR: 17 to 31%) of exposures across pairwise combinations of sixteen common antibiotics and nine bacterial pathogens. All scenarios demonstrate that stewardship interventions, facilitated by changes in clinician behavior and improved diagnostics, have the opportunity to broadly reduce antibiotic exposures across a range of potential pathogens. Concurrent approaches, such as vaccines aiming to reduce infection incidence, are needed to further decrease exposures occurring in “necessary” contexts.

## Introduction

Antibiotic consumption is a known driver of antibiotic resistance. In developed nations, over 80% of antibiotic consumption for human health occurs in the outpatient setting (European Centre for Disease Prevention and Control, 2018; Public Health Agency of Canada, 2018; Public Health England, 2017; Swedres-Svarm, 2017), and US-based studies conducted across different subpopulations have estimated that 23-40% of outpatient prescriptions may be inappropriate (Chua, Fischer, & Linder, 2019; Fleming-Dutra et al., 2016; Olesen, Barnett, MacFadden, Lipsitch, & Grad, 2018). Inappropriate antibiotic use leads to increased risk of adverse events (Linder, 2008; Shehab, Patel, Srinivasan, & Budnitz, 2008), disruption of colonization resistance and other benefits of the microbial flora (Buffie & Pamer, 2013; Khosravi & Mazmanian, 2013), and bystander selection for antibiotic resistance, with little to no health gains for the patient.

Recent work on bystander selection estimates that, for 8 of 9 potential pathogens of interest, over 80% of their exposures to commonly used antibiotic classes in the outpatient setting occur when the organisms are asymptomatically colonizing the microbiome, not causing disease (Tedijanto, Olesen, Grad, & Lipsitch, 2018). A corollary of extensive bystander selection is that reductions in use will prevent antibiotic exposures for species throughout the microbiome.

We use publicly available data to quantify potentially avertable exposures of bacterial species to commonly used antibiotics under hypothetical changes in prescribing practice. The set of scenarios included here are intended as thought experiments to explore the upper bounds of avertable antibiotic exposures. Reductions in antibiotic consumption have historically had variable impacts on resistance levels, likely dependent on setting, baseline consumption and resistance patterns, and fitness costs (Andersson & Hughes, 2010; Lipsitch, 2001). In addition, treatment strategies are primarily guided by randomized controlled trials assessing immediate clinical outcomes, with less careful consideration given to the microbiome-wide effect of such decisions. We apply analytical methods to parse out species-level effects of changes in prescribing practice in order to better understand the potential impact of antibiotic stewardship.

## Methods

### Scenarios of interest

This analysis includes sixteen antibiotics that are frequently prescribed in the outpatient setting and nine potentially pathogenic bacterial species that are commonly carried in the normal human microbiome. For each antibiotic-species pair, we estimate the proportion of antibiotic exposures experienced by that species that could be averted under four hypothetical scenarios. The scenarios range from broad elimination of unnecessary prescribing to focused modifications of antibiotic use for specific indications as follows:

1. Eliminate unnecessary antibiotic use across all outpatient conditions.
2. Eliminate all antibiotic use for outpatient respiratory conditions for which antibiotics are not indicated.
3. Eliminate all antibiotic use for acute sinusitis.
4. Prescribe nitrofurantoin for all cases of cystitis in women.

We use the results reported by Fleming-Dutra and colleagues as estimates of unnecessary antibiotic use in ambulatory care settings (Fleming-Dutra et al., 2016). In this study, the authors convened a group of experts to estimate the proportion of appropriate antibiotic use by condition and age group based on clinical guidelines. When guidelines could not be used for this task, the rate of appropriate antibiotic prescribing was estimated by benchmarking against the lowest-prescribing US region. Based on the same paper, we consider viral upper respiratory tract infection, influenza, non-suppurative otitis media, viral pneumonia, bronchitis, and allergy and asthma to be outpatient respiratory conditions for which antibiotics are not indicated.

Antibiotic treatment of acute bacterial sinusitis is currently guideline-recommended (Rosenfeld et al., 2015). However, bacteria are an infrequent cause of acute sinusitis, and due to the self-limiting nature of the syndrome, evidence to support antibiotic treatment is weak (Burgstaller, Steurer, Holzmann, Geiges, & Soyka, 2016; Fokkens, Lund, & Mullol, 2007). The cause of sinusitis (whether bacterial, viral, or noninfectious) can be very difficult to distinguish in practice, and as a result of this challenge and others, antibiotics continue to be prescribed at over 80% of US outpatient visits with a primary diagnosis of acute sinusitis (Smith et al., 2013). In contrast, antibiotics are always recommended for urinary tract infections (UTIs). Despite being recommended as a second-line therapy for cystitis due to concerns about resistance, fluoroquinolones are the most common treatment, prescribed at 40% of outpatient visits for uncomplicated UTI (Kabbani et al., 2018). We explore the hypothetical scenario of treating all cases of cystitis in women with nitrofurantoin, a recommended first-line therapy with good potency against common uropathogens, low levels of resistance, and decreased risk of collateral damage to the intestinal microbiome due to its propensity to concentrate in the bladder (Gupta et al., 2011; Gupta, Scholes, & Stamm, 1999; Stewardson et al., 2015). Nitrofurantoin is considered clinically appropriate unless the patient has chronic kidney disease, is showing signs of early pyelonephritis, or has had a prior urinary isolate resistant to nitrofurantoin (Hooton & Gupta, 2019). In our analysis, we exclude patients with a simultaneous diagnosis of pyelonephritis. We assume that other contraindications are fairly rare in the general population.

### Data sources and methodology

Extending our recent work on bystander selection (Tedijanto et al., 2018), we used the 2015 National Ambulatory Medical Care Survey and National Hospital Ambulatory Medical Care Survey (NAMCS/NHAMCS) (National Center for Health Statistics, 2015a, 2015b), the Human Microbiome Project (HMP) (Huttenhower et al., 2012; The Human Microbiome Project Consortium, 2012), and assorted carriage and etiology studies (Bäckhed et al., 2015; Bluestone, Stephenson, & Martin, 1992; Bogaert et al., 2011; Brook, Thompson, & Frazier, 1994; Celin, Bluestone, Stephenson, Yilmaz, & Collins, 1991; Chira & Miller, 2010; Edlin, Shapiro, Hersh, & Copp, 2013; Ginsburg et al., 1985; Gunnarsson, Holm, & Söderström, 1997; Gupta et al., 1999; Jack M. Gwaltney, Scheld, Sande, & Sydnor, 1992; Hammitt et al., 2006; Holgerson, Öhman, Rönnlund, & Johansson, 2015; Susan S. Huang et al., 2009; Jain et al., 2015; Arch G. Mainous, Hueston, Everett, & Diaz, 2006; Pettigrew et al., 2012; Regev-Yochay et al., 2004; Verhaegh et al., 2010; Loretta Wubbel et al., 1999; Yassour et al., 2016) (details in Figure 1 – Source Data File 1) to estimate national outpatient antibiotic exposures by drug, species, and condition.

NAMCS/NHAMCS are annual cross-sectional surveys designed to sample outpatient visits in the United States, and up to five diagnoses and thirty medications may be associated with each visit. We used methodology developed by Fleming-Dutra and colleagues (Fleming-Dutra et al., 2016), and applied in other studies (Olesen, Barnett, MacFadden, Lipsitch, et al., 2018), to group diagnosis codes into conditions and link antibiotic prescriptions with the most likely indication. For this analysis, visits with a diagnosis of acute cystitis (ICD-9-CM: 595.0), unspecified cystitis (595.9) or unspecified UTI (599.0), without a concurrent diagnosis of pyelonephritis (590.1, 590.8), renal abscess (590.2), or kidney infection (590.9), were considered to be associated with cystitis. In this analysis, we maintain the assumption that one antibiotic prescription is equivalent to one exposure; antibiotic exposures experienced by a given species and associated with a given condition are roughly estimated as the product of antibiotic prescriptions for that condition and species carriage prevalence, which is dependent on disease etiology (target exposures) and asymptomatic carriage prevalence (bystander exposures). For diagnoses where etiology was not readily available, we assumed that none of our species of interest were causative agents. These assumptions have been enumerated in detail in prior work (Tedijanto et al., 2018). We applied proportions of unnecessary antibiotic prescribing by condition and age group estimated by Fleming-Dutra and colleagues based on expert opinion, clinical guidelines, and regional variability in use (Fleming-Dutra et al., 2016). For relevant scenarios (1 and 2), we applied the proportions of unnecessary use evenly across all antibiotic prescriptions. Antibiotics and antibiotic classes are identified by the Lexicon Plus® classification scheme (*Lexicon Plus*, 2008).

The proportion of avertable antibiotic exposures for each species is defined by Equation 1. The equation adopts previously described notation (Tedijanto et al., 2018) with modifications. A listing of all variables and descriptions can be found in Equation 1 – Figure Supplement 1. Let *a* represent antibiotic, *s* represent species, *i* represent ICD-9-CM diagnosis code, and *g* represent age group. Throughout the analysis we have weighted outpatient visits to be nationally representative using the sampling and nonresponse weights provided in NAMCS/NHAMCS. Let *X*_*as*_ be the number of avertable exposures by antibiotic and species, *T*_*as*_ be the total number of exposures by antibiotic and species, *d*_*aig*_ be the number of prescriptions of antibiotic *a* associated with diagnosis code *i* in age group *g, p*_*sig*_ be the carriage prevalence of species *s* among those with diagnosis code *i* in age group *g*, and *q*_*aig*_ be the proportion of avertable exposures by diagnosis code and age group in the given scenario. For example, in the scenario assessing elimination of all unnecessary antibiotic use, *q*_*aig*_ is the proportion of avertable antibiotic use by diagnosis and age group (Fleming-Dutra et al., 2016). Alternatively, in the scenario assessing elimination of non-nitrofurantoin treatment for cystitis, *q*_*aig*_ is 1 when *a* is not nitrofurantoin, *i* is a diagnosis code associated with cystitis, and the patient is female, and 0 elsewhere. Carriage prevalences (*p*_*sig*_) are assumed to be constant within three age groups (under 1 year, 1-5 years, over 5 years old) (Tedijanto et al., 2018), while proportions of avertable antibiotic use (*q*_*aig*_) were reported for three different age groups (0-19 years, 20-64 years, 65 years old and over) (Fleming-Dutra et al., 2016). *G* is the smallest set of age groups that accounts for this granularity (under 1 year, 1-5 years, 6-19 years, 20-64 years, 65 years old and over). For antibiotic prescriptions that occurred at visits without any ICD-9-CM diagnosis codes (i=0), we applied the carriage prevalence among healthy individuals.

**Equation 1**. Proportion of avertable exposures by species and antibiotic. Equation 1 – Figure Supplement 1 includes further details on all variables.

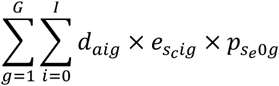

In scenarios where unnecessary use is eliminated (1 and 2), we assume that only bystander exposures are affected. Equivalently, this presumes that perfect discrimination between bacterial and non-bacterial etiologies is possible. This results in a slight modification to the numerator of Equation 1 -- *p*_*sig*_ is changed to *p*_*s0g*_ as all eliminated exposures would have occurred during asymptomatic carriage. For cases where *e*_*sig*_ is 1, we assume zero avertable exposures. In addition, we make slight modifications to *q*_*aig*_ when we estimate the proportion of bacterial cases to be larger than the proportion of necessary prescriptions (Figure 1 – Source Data File 2) (Bluestone et al., 1992; Brook, 2016; Celin et al., 1991). As a sensitivity analysis, we include the proportion of avertable exposures for each bacteria-antibiotic pair under Scenario 1 if antibiotic use was eliminated equally across both target and bystander exposures (Figure 1 – Figure Supplement 1). All analysis was conducted in R version 3.6.1.

## Results

Results for all four scenarios are depicted in Figure 1.

**Figure 1.**
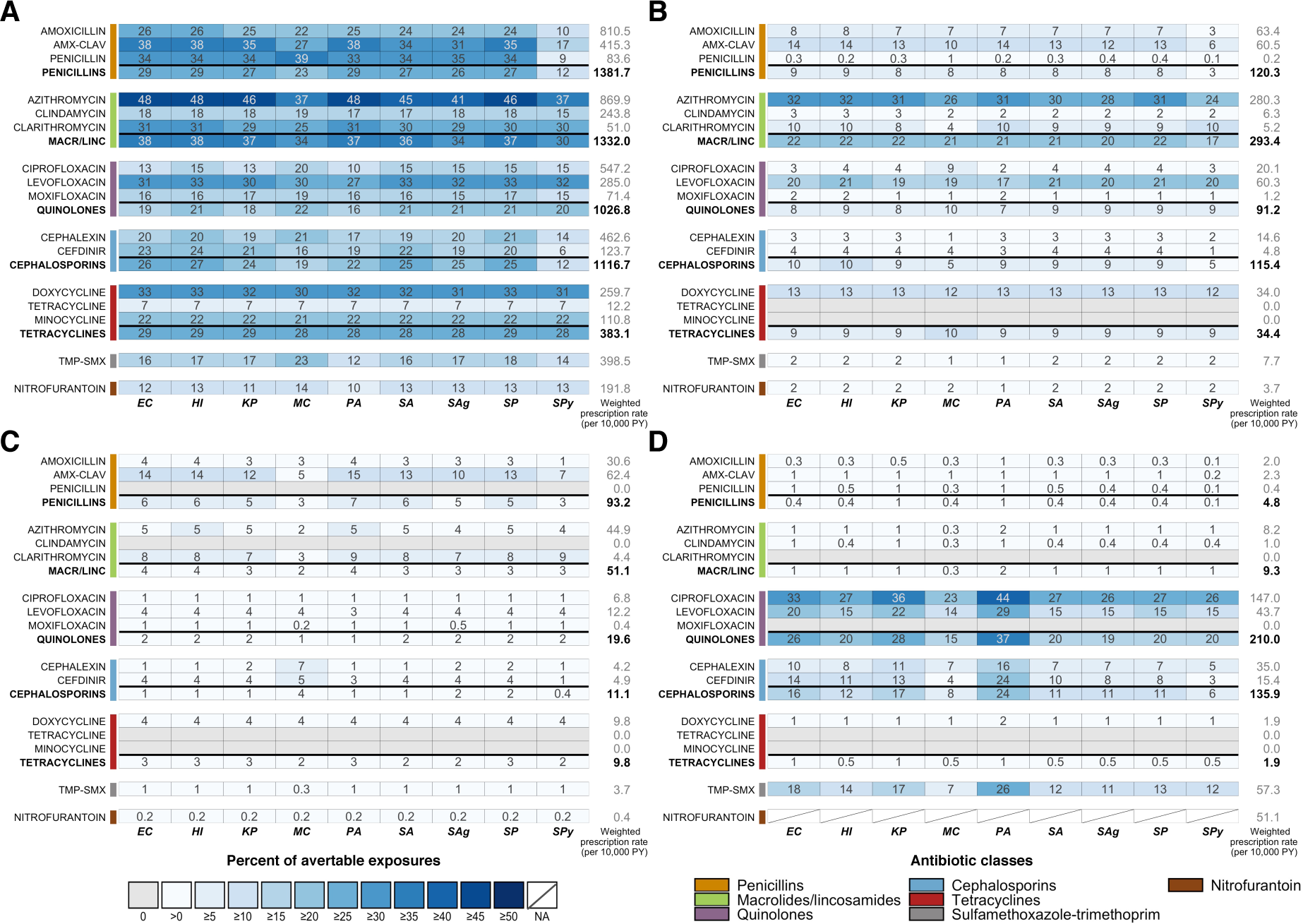
Heatmaps showing the estimated percentage of species exposures to each antibiotic or antibiotic class that could be averted if *A:* unnecessary antibiotic prescriptions across all outpatient conditions, *B:* all antibiotic use for outpatient respiratory conditions for which antibiotics are not indicated, *C:* all antibiotic use for acute sinusitis, or *D:* non-nitrofurantoin treatment of cystitis in women was eliminated. Drug class results include prescriptions of all antibiotics in that class, as identified by the Lexicon Plus® classification system. Sensitivity and other additional analyses are shown in Figure 1 – Figure Supplements 1-4. **Abbreviations:** *Antibiotics (y-axis):* AMX-CLAV=amoxicillin-clavulanate,, MACR/LINC=macrolides/ lincosamides, TMP-SMX=sulfamethoxazole-trimethoprim; *Organisms (x-axis):* EC=*E. coli*, HI=*H. influenzae*, KP=*K. pneumoniae*, MC=*M. catarrhalis*, PA=*P. aeruginosa*, SA=*S. aureus*, SAg=*S. agalactiae*, SP=*S. pneumoniae*, SPy=*S. pyogenes*; PY=person-years

### Scenario 1

We estimate that elimination of unnecessary antibiotic prescriptions across all outpatient conditions would prevent 6 to 48% (IQR: 17 to 31%) of antibiotic-species exposures (Figure 1A). The smallest reduction is associated with *S. pyogenes* exposures to cefdinir and the largest with *H. influenzae, E. coli*, and *P. aeruginosa* exposures to azithromycin. If all unnecessary antibiotic use could be prevented, over 30% of exposures to amoxicillin-clavulanate, penicillin, azithromycin, clarithromycin, levofloxacin, and doxycycline across most potential pathogens of interest could be averted. Of particular interest, elimination of all unnecessary prescribing in the outpatient setting could reduce exposures of *S. pneumoniae* to penicillins and macrolides by 27% and 37%, respectively, and of *S. aureus* to penicillins and quinolones by 27% and 21%. For *E. coli* and *K. pneumoniae*, approximately one-quarter of exposures to cephalosporins and one-fifth of exposures to quinolones could be averted.

### Scenario 2

Scenario 2, elimination of all antibiotic use for outpatient respiratory conditions where antibiotics are not indicated, is a subset of Scenario 1 and accounts for a substantial portion of avertable exposures included in the first scenario. As in Scenario 1, the results are primarily driven by drug and depend on the amount of use of that drug for the affected conditions (in this case, conditions for which antibiotics are not indicated). Pathogen characteristics affecting the proportion of avertable exposures can be better understood by looking across all species for a single drug. For example, we focus on the row for azithromycin. Based on NAMCS/NHAMCS 2015, 32% of all azithromycin use is associated with the affected conditions. Since these conditions are never caused by our organisms of interest, the maximum proportion of avertable exposures for any species is 32%. However, the proportion of avertable exposures is modulated by the number of azithromycin exposures that occur for treatment of other conditions – for example, a lower proportion of avertable exposures for *S. pyogenes, M. catarrhalis*, and *S. agalactiae* indicates that they have relatively frequent exposure to azithromycin during treatment of other conditions that are unaffected in this scenario, often both as a bystander and causative pathogen.

### Scenario 3

In Scenario 3, we explore elimination of all antibiotic use for acute sinusitis. Overall, acute sinusitis accounts for just 3% of prescriptions of our included antibiotic classes, resulting in the low proportions of avertable exposures across most bacteria-antibiotic combinations. However, proportions of avertable exposures are relatively high for amoxicillin-clavulanate and clarithromycin, antibiotics for which a large proportion of their use is due to acute sinusitis (15% and 9%, respectively). In this scenario, we make the assumption that all use for acute sinusitis is unnecessary. In reality, a small number of cases are truly bacterial and antibiotics are indicated for patients with persistent, severe or worsening symptoms (Rosenfeld et al., 2015). We explore varying levels of unnecessary use based on etiological data in Figure 1 – Figure Supplement 2 (Aitken & Taylor, 1998; Benninger, Holzer, & Lau, 2000; Fleming-Dutra et al., 2016; Sande & Gwaltney, 2004; Snow, Mottur-Pilson, & Hickner, 2001; Wald, Guerra, & Byers, 1991; Williams, Simel, Roberts, & Samsa, 1992).

### Scenario 4

In this scenario, we consider a program to prescribe only nitrofurantoin for all cases of acute cystitis in women, replacing all other antibiotics that are currently in use. As a result of this change, we estimate that exposures of *E. coli* and *K. pneumoniae* to cephalosporins could be reduced by 16% and 17%, respectively, and exposures to quinolones could decrease by 26% and 28%. Uniquely in this scenario among the ones we considered, several of our species of interest cause a large proportion of UTI cases (*E. coli, K. pneumoniae*, and *P. aeruginosa*). To explore the effects of being a causative pathogen on the proportion of avertable exposures, we focus on the results for ciprofloxacin. Overall, 27% of ciprofloxacin use is associated with acute cystitis in women. Organisms that are not causative pathogens of cystitis have proportions of avertable exposures close to or less than 27%, but a substantially higher proportion of exposures could be averted among causative pathogens - 33%, 36%, and 44% of ciprofloxacin exposures to *E. coli, K. pneumoniae*, and *P. aeruginosa*, respectively. Although *E. coli* is the most common cause of UTIs, its estimated proportion of avertable exposures in this scenario is lower than that of *P. aeruginosa*, likely due to the larger number of antibiotic exposures incurred by *E. coli* during asymptomatic carriage.

This scenario also allows us to observe the impact of differences in disease incidence, antibiotic use patterns, and carriage prevalence across age groups on the proportion of avertable exposures. For example, *M. catarrhalis* is asymptomatically carried in children much more frequently than in adults. Since cystitis occurs primarily in adults, the proportion of avertable exposures is often lower for *M. catarrhalis* compared to other species due to bystander exposures in children that are largely unaffected by modified prescribing practices for cystitis. This age group effect can be substantial for antibiotics that are commonly given to children, especially when the antibiotic is also used to treat a condition caused by *M. catarrhalis* (e.g. cefdinir and otitis media), but is less influential for antibiotics that are infrequently prescribed to children, such as ciprofloxacin and levofloxacin.

Overall, the proportion of avertable exposures is driven by pathogen, syndrome, and prescribing characteristics. Because our organisms of interest experience the vast majority of their antibiotic exposures as bystanders, the metric is largely dependent on the proportion of antibiotic use that is associated with the conditions affected in the given scenario. In a scenario where all antibiotic use for affected conditions is eliminated, this proportion would be equal to the proportion of avertable exposures for organisms exposed to antibiotics only during carriage (never pathogens). Being a causative pathogen for any of the affected conditions increases the proportion of avertable exposures. In contrast, being a causative pathogen for unaffected conditions increases unaffected antibiotic exposures and thus decreases the proportion of avertable exposures. For pathogens responsible for multiple syndromes, both factors may be at play. Finally, asymptomatic carriage prevalence is positively associated with both affected and unaffected antibiotic exposures, and, as a result, its overall effect on the proportion of avertable exposures is dependent on the bacteria-antibiotic combination of interest.

## Discussion

We quantify the species-level impact of changes in antibiotic consumption as the proportion of antibiotic exposures experienced by common bacterial pathogens that could be averted under four hypothetical scenarios. In the scenario where unnecessary antibiotic use for all outpatient conditions is eliminated, we find that up to 48% of exposures (of *H. influenzae, E. coli*, and *P. aeruginosa* to azithromycin) could be avoided. In addition, impact of the intervention across antibiotics and species is widespread, with half of antibiotic-species pairs expected to experience a reduction in exposures of over one-fifth (22%). For conditions which require antibiotic treatment, such as UTIs, switching to antibiotics with decreased collateral damage to the microbiome, such as nitrofurantoin, may be an effective strategy for reducing antibiotic exposures across species.

In three out of four situations we assess, antibiotics are considered entirely unnecessary for some or all cases. In the future, this method may also be extended to measure net changes in exposures resulting from more nuanced scenarios where one antibiotic is substituted for another. We did not assess such changes for nitrofurantoin, as its effects on the microbiome outside of the bladder are thought to be minimal (Stewardson et al., 2015). The reductions in use presented here may be practically infeasible due to challenges including similar clinical presentation of viral and bacterial infections, laboratory processing times that prevent identification of causative pathogens during outpatient visits, individual considerations such as allergies (Sakoulas, Geriak, & Nizet, 2018) or heightened risk of adverse events, misdiagnosis (Filice et al., 2015; Tomas, Getman, Donskey, & Hecker, 2015), and patient-driven demand (Vanden Eng et al., 2003). Even if elimination of unnecessary use were fully realized, our results imply that the majority of species’ antibiotic exposures occur in the context of “necessary” antibiotic use. These findings underscore the importance of considering bystander effects and the need for a multi-pronged approach to programs aimed at controlling antibiotic resistance. Rapid diagnostics, diagnostics that accurately discriminate between bacterial and non-bacterial causes, patient education, and improved decision-making tools and other interventions to motivate changes in clinician behavior can enhance responsible antibiotic consumption and reduce unnecessary antibiotic use; these should be implemented simultaneously with prevention measures such as infection control, access to clean water and sanitation, safe sex interventions, and vaccination, which aim to reduce infection incidence and thus any antibiotic use.

At the time of this analysis, NHAMCS data from hospital outpatient departments was not available for 2015. In the 2010-2011 NAMCS/NHAMCS data, visits to hospital outpatient departments accounted for 36% of included sampled visits, but just 9% of all antibiotic mentions. Since the proportions of avertable exposures including and excluding these visits for 2010-2011 were similar for Scenario 1 (Figure 1 – Figure Supplement 3), we assumed that the existing 2015 data without hospital outpatient department visits was representative of the NAMCS/NHAMCS population for the purposes of our analysis. Certain outpatient encounters are also outside the scope of NAMCS/NHAMCS, including federal facility visits, telephone contacts, house calls, long-term care stays, urgent care, retail clinics, and hospital discharge prescriptions.

Additional limitations of the included datasets and method for estimating antibiotic exposures have been previously enumerated (Tedijanto et al., 2018). Notably, the NAMCS/NHAMCS does not provide information to link medications with diagnoses, so we adopted a published tiered system to pair antibiotic use at each visit with the most likely indication (Fleming-Dutra et al., 2016). For visits with multiple diagnoses, all antibiotic use was attributed to the single most-likely indication. This method tends to overestimate antibiotic use for conditions for which antibiotics are almost always indicated, which may lead to clinically unusual diagnosis-treatment linkages. In the same way, antibiotic prescriptions are underestimated for diagnoses for which antibiotic use is not indicated, potentially leading to downward bias in avertable exposures for these conditions. Additionally, NAMCS/NHAMCS antibiotic use data does not include prescription details such as duration or dose. Future studies with more granular data may extend the methods presented here to account for such factors.

At the time of this analysis, Fleming-Dutra et al. remained the most up-to-date source of unnecessary antibiotic prescribing by condition. Recent work with slightly different methods found a lower proportion of inappropriate antibiotic prescriptions overall (23.2% compared to 30%) but did not report their estimates by condition (Chua et al., 2019). Although comparable estimates of unnecessary prescribing over time are unavailable, multiple studies have shown that declines in antibiotic use from approximately 2011 to 2016 in the United States have been primarily due to pediatric prescribing, implying that, at least for adults, levels of unnecessary prescribing likely remained similar (Durkin et al., 2018; King, Bartoces, Fleming-Dutra, Roberts, & Hicks, 2019; Klevens et al., 2019; Olesen, Barnett, MacFadden, Lipsitch, et al., 2018). We also assume that the proportion of unnecessary use is constant across all antibiotic use for the same condition as it is difficult to identify specific antibiotic prescriptions that were unwarranted without detailed chart review. A study among outpatients in the Veterans Affairs medical system found that among prescriptions for community-acquired pneumonia, sinusitis, and acute exacerbations of chronic bronchitis, the highest proportion of macrolide use was inappropriate (27%), followed by penicillins (22%) and quinolones (12%) (Tobia, Aspinall, Good, Fine, & Hanlon, 2008). Similar studies are needed to understand which antibiotics are frequently used inappropriately for other indications and settings.

Finally, it is important to note that the reduction in antibiotic exposures estimated here does not translate to the same reduction in the prevalence of resistance or in the morbidity and mortality attributable to resistance. Although population-level antibiotic consumption has been positively correlated with levels of antibiotic resistance, the impact of changing consumption is not well-understood and is likely to vary widely by bacteria-antibiotic combination (or even strain-antibiotic combination) based on resistance mechanisms, fitness costs, and co-selection, among other factors (Andersson & Hughes, 2010; Pouwels et al., 2017). Antibiotic consumption is also highly heterogeneous and the impact of stewardship on resistance may depend on the affected patient populations (Olesen, Barnett, MacFadden, Brownstein, et al., 2018). Additionally, in this analysis, we give each exposure equal weight. However, the selective pressure imposed by a single exposure depends on a number of variables, including pharmacokinetics, pharmacodynamics, distribution of bacteria across body sites, and bacterial population size. For example, we might expect that the probability of resistance scales with population size, and thus that an exposure received by an individual with higher bacterial load will have a larger impact on resistance. Further research, integrating knowledge from clinical, ecological, and evolutionary spheres, is needed to elucidate the relationship between antibiotic use and selective pressures and ultimately between use and resistance at the population level (MacLean & San Millan, 2019).

Reductions in antibiotic consumption are necessary to preserve the potency of these drugs. Quantifying changes in species-level exposures due to stewardship programs is one more step towards understanding how changes in antibiotic use correspond to antibiotic resistance. The methods presented here may be easily extended to incorporate other data sources, such as claims, or to assess more specific stewardship programs. We show that while improved prescribing practices have the potential to prevent antibiotic exposures experienced by bacterial species throughout the microbiome, complementary efforts to facilitate appropriate antibiotic consumption and decrease overall infection incidence are required to substantially avert exposures.

## Data Availability

Data is publicly available.

https://www.cdc.gov/nchs/ahcd/datasets_documentation_related.htm

https://www.hmpdacc.org/resources/data_browser.php

## Acknowledgements

We thank Dr. Lauri Hicks for her helpful comments on this manuscript.

## Competing Interests

ML has received consulting income from Affinivax, Antigen Discovery, Merck, and Pfizer and research grants through Harvard School of Public Health from Pfizer and PATH. YHG has received consulting income from Merck and GlaxoSmithKline. The authors have no other relationships or activities that could appear to have influenced the submitted work.

## Funding Sources

This work was supported by Grant U54GM088558 (Models of Infectious Disease Agent Study, Center for Communicable Disease Dynamics) from the National Institute of General Medical Sciences, Grant R01AI132606 from the National Institute of Allergy and Infectious Diseases (to YHG), Grant CK000538-01 from the Centers for Disease Control and Prevention, and the Doris Duke Charitable Foundation. CT is supported by Grant T32AI007535 from the National Institute of Allergy and Infectious Diseases. The content is solely the responsibility of the authors and does not necessarily represent the official views of the National Institute of General Medical Sciences, National Institutes of Health, National Institute of Allergy and Infectious Diseases, Centers for Disease Control and Prevention, Department of Health and Human Services, or the Doris Duke Charitable Foundation.

## Figure Supplements

*Equation 1 – Figure Supplement 1*. Notation, descriptions, and sources for variables in Equation 1.

*Figure 1 – Figure Supplement 1*. Sensitivity analysis of proportions of avertable exposures across all outpatient conditions when proportion of unnecessary use is applied equally across target and bystander exposures (Equation 1 without modification).

*Figure 1 – Figure Supplement 2*. Sensitivity analyses for Scenario 3 (elimination of all antibiotic use for acute sinusitis).

*Figure 1 – Figure Supplement 3*. Proportions of avertable exposures under Scenario 1 (elimination of all unnecessary antibiotic use across outpatient conditions) using 2010-2011 NAMCS/NHAMCS data with and without NHAMCS outpatient department data.

## Source Data Files

*Figure 1 – Source Data File 1*. Summary of etiology studies.

*Figure 1 – Source Data File 2*. Modifications to proportions of unnecessary antibiotic prescriptions based on published etiology studies and antibiotic use in NAMCS/NHAMCS. Modifications were made if the reported proportion of bacterial cases for a given condition exceeded the estimated proportion of appropriate prescriptions reported in Fleming-Dutra et al.

*Figure 1 – Source Data File 3*. Estimated antibiotic class exposures by exposed and causative species.

---

**Equation 1 – Figure Supplement 1.**
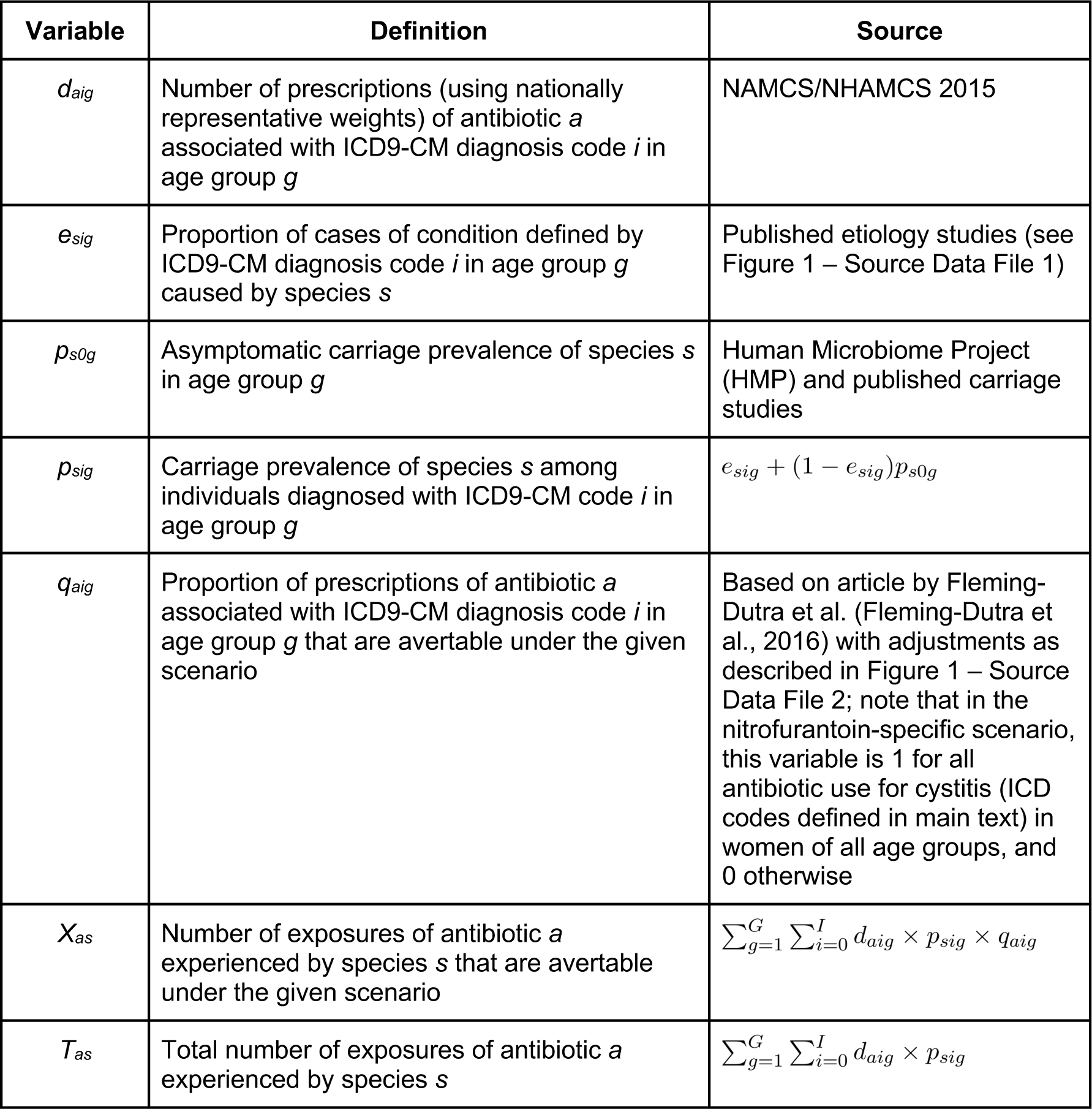
Notation, descriptions, and sources for variables in Equation 1.

**Figure 1 – Figure Supplement 1.**
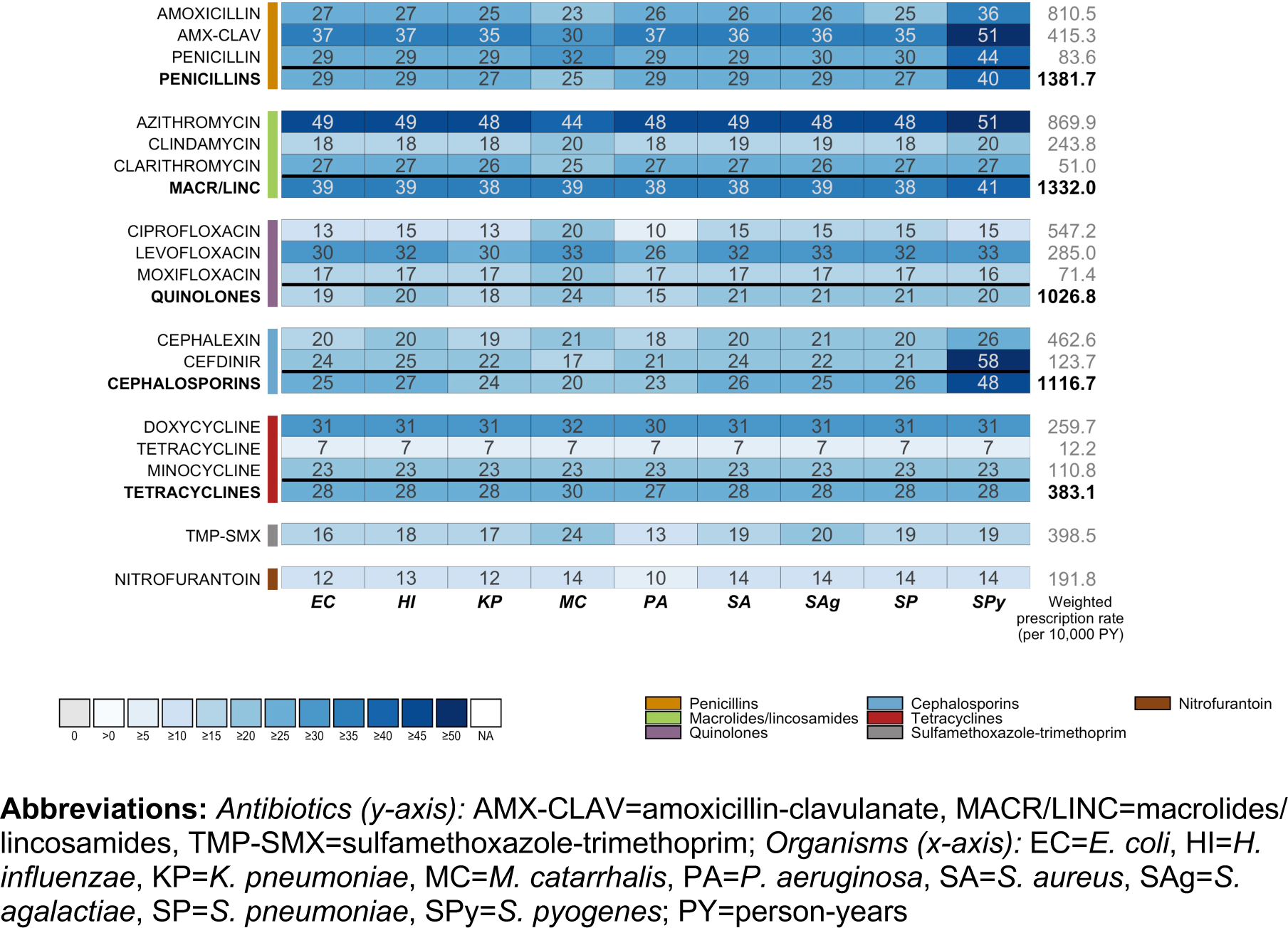
Sensitivity analysis of proportions of avertable exposures across all outpatient conditions when proportion of unnecessary use is applied equally across target and bystander exposures (Equation 1 without modification).

**Figure 1 – Figure Supplement 2.**
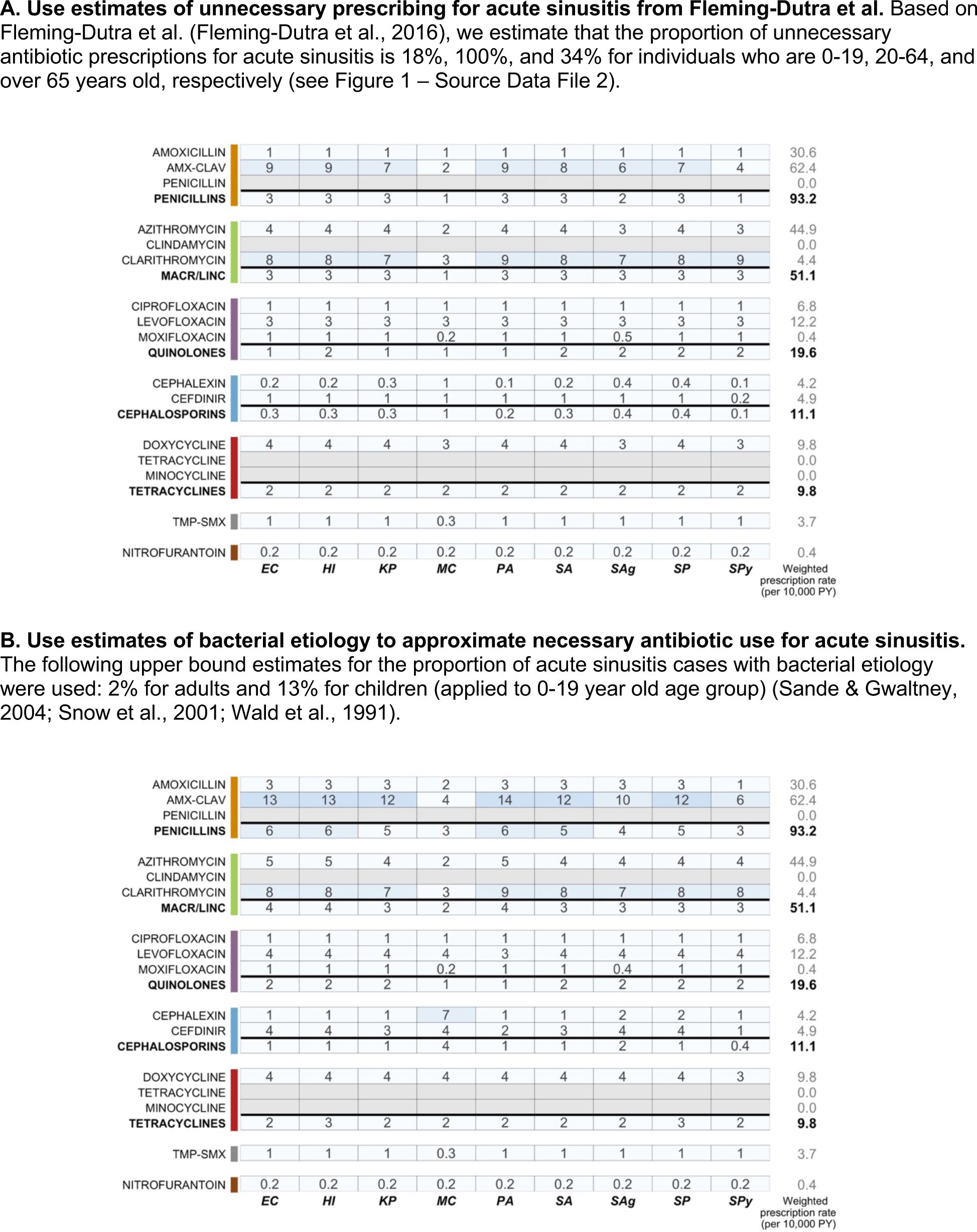
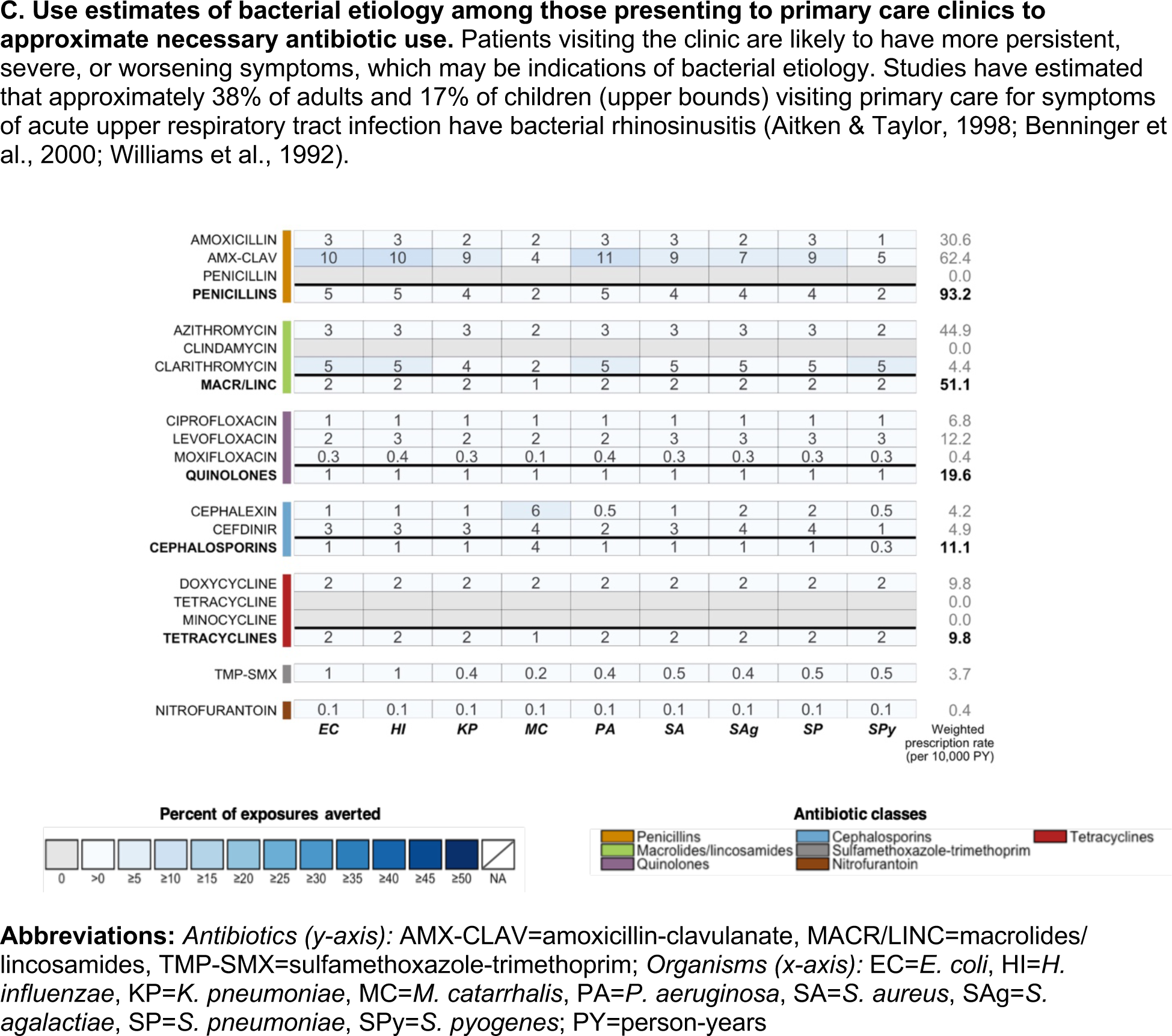
Sensitivity analyses for Scenario 3 (elimination of all antibiotic use for acute sinusitis).

**Figure 1 – Figure Supplement 3.**
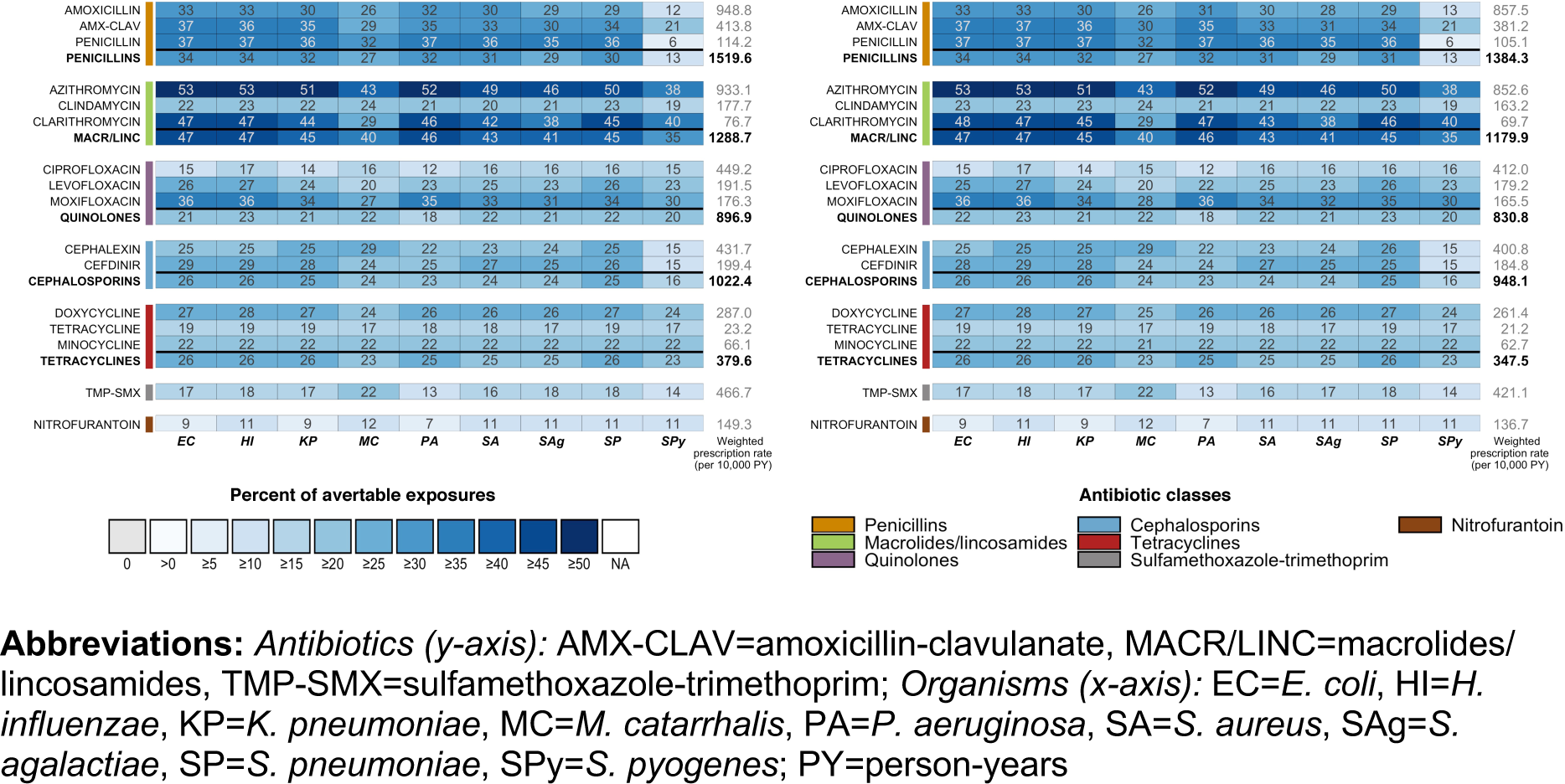
Proportions of avertable exposures under Scenario 1 (elimination of all unnecessary antibiotic use across outpatient conditions) using 2010-2011 NAMCS/NHAMCS data with *(left)* and without *(right)* NHAMCS outpatient department data.

**Figure 1 – Source Data File 1.**
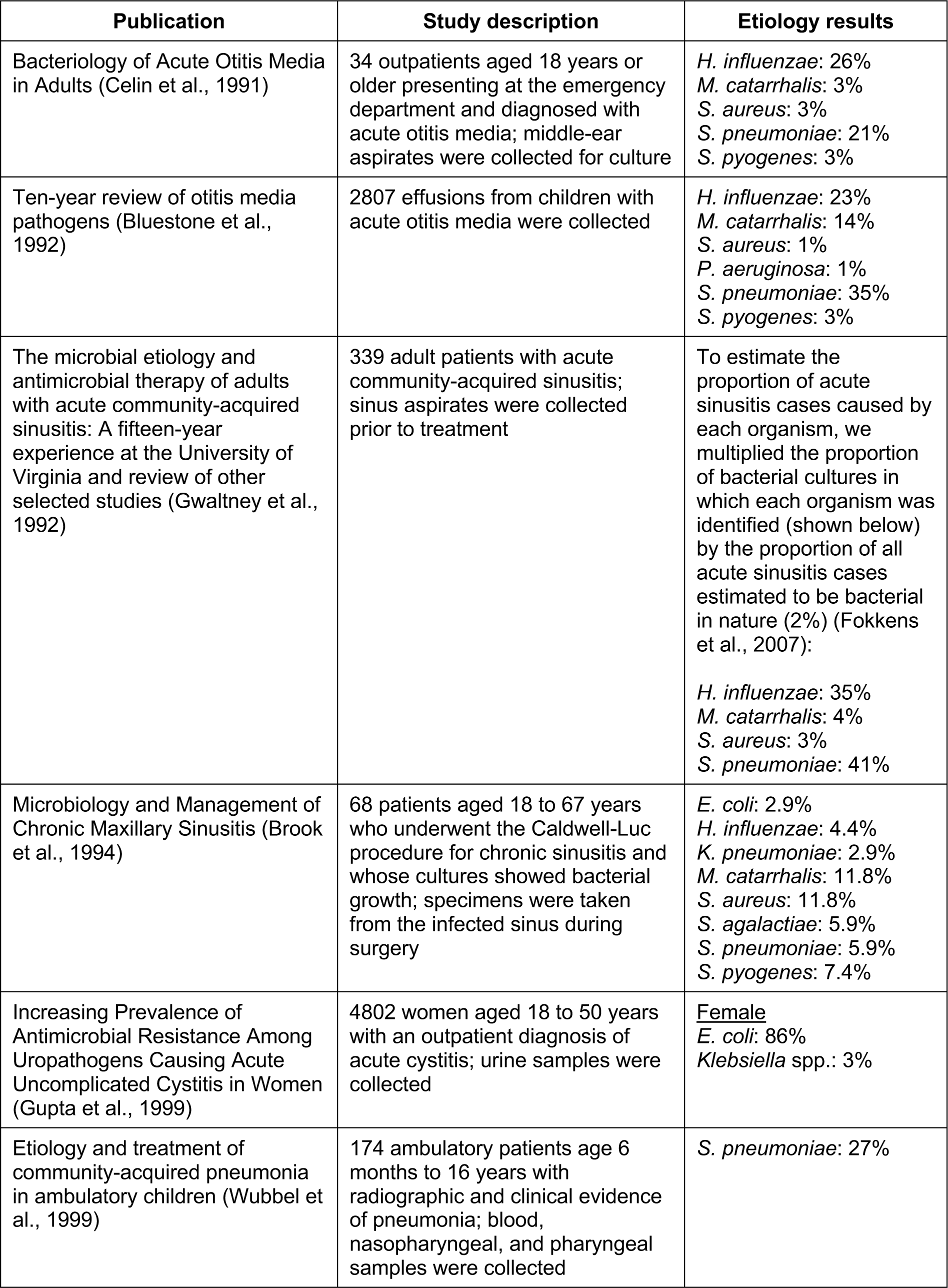
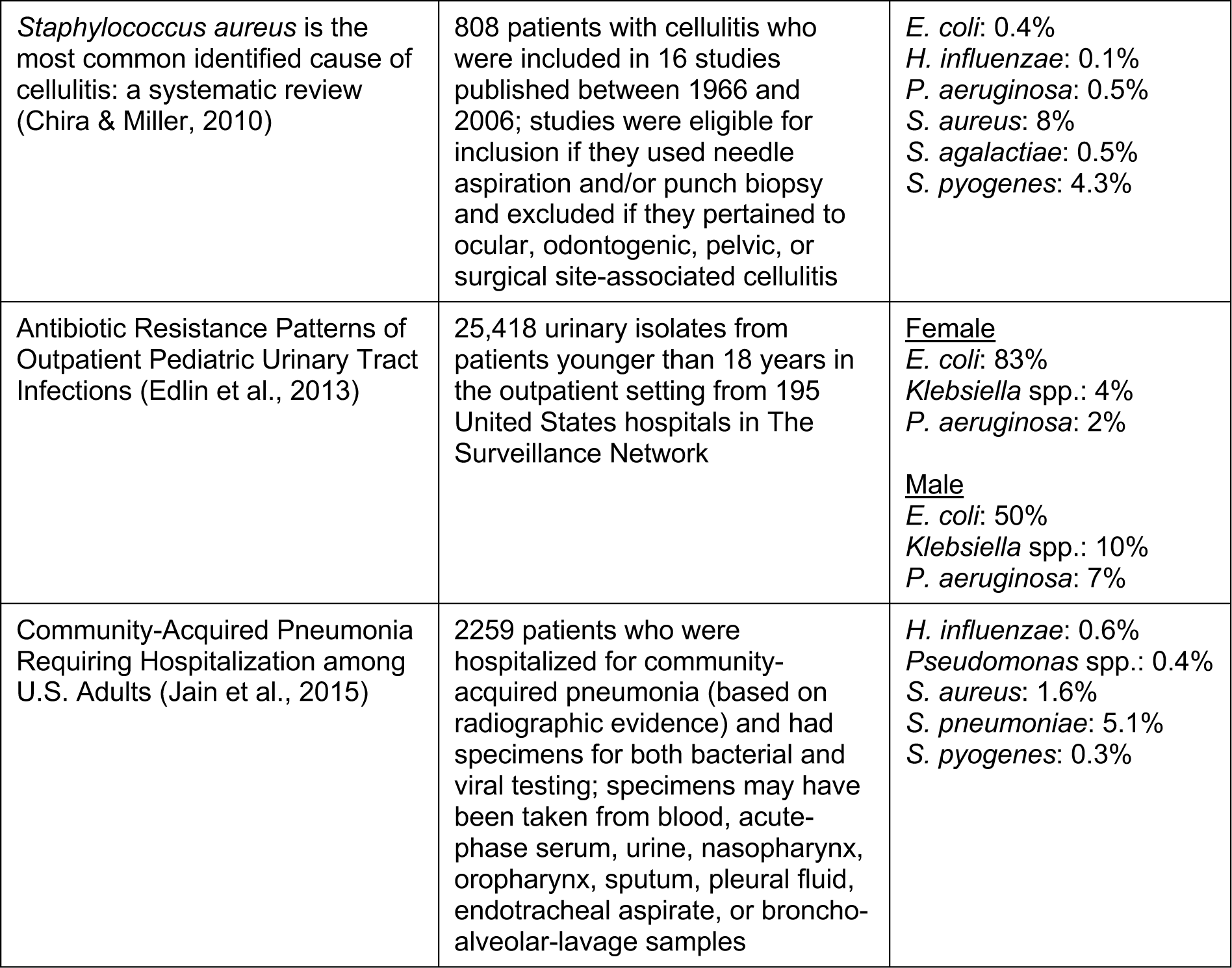
Summary of etiology studies.

**Figure 1 – Source Data File 2.**
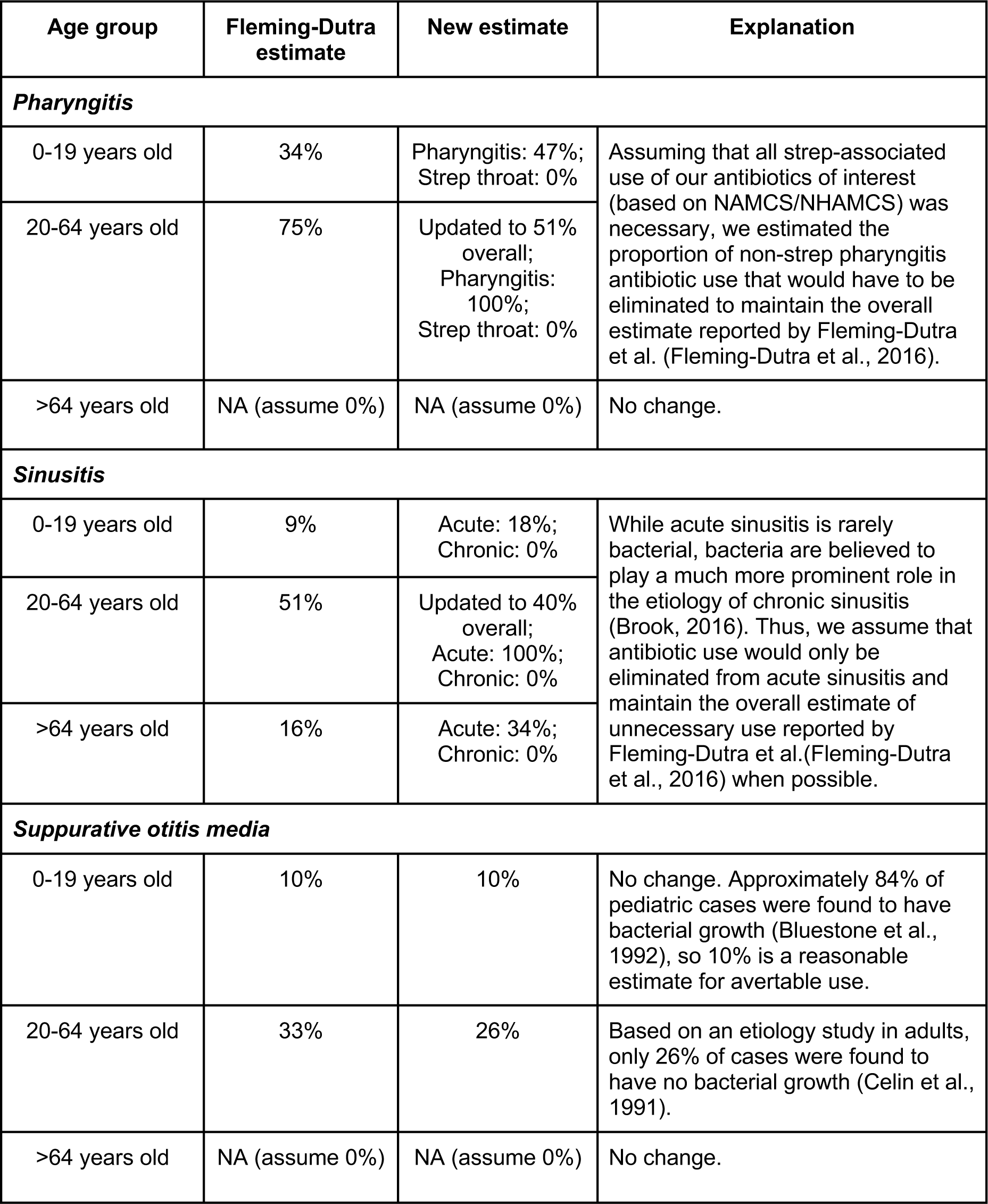
Modifications to proportions of unnecessary antibiotic prescriptions based on published etiology studies and antibiotic use in NAMCS/NHAMCS. Modifications were made if the estimated proportion of bacterial cases for a given condition exceeded the estimated proportion of appropriate prescriptions reported in Fleming-Dutra et al. (Fleming-Dutra et al., 2016).

## Figure 1 – Source Data File 3.

Estimated antibiotic class exposures by exposed and causative species.

The following tables contain the estimated number of annual antibiotic exposures (in 100,000s) in the outpatient setting in the United States by the species being exposed (y-axis) and the species that is the cause of antibiotic use (x-axis). The values in each table, excluding the “Other” and “Overall” columns were calculated using the following expression:

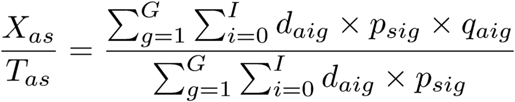

Let *a* represent antibiotic class, *s*_*c*_ represent causative species, *s*_*e*_ represent exposed species, *i* represent ICD-9-CM diagnosis code, and *g* represent age group (under 1 year, 1-5 years, 6-19 years, 20-64 years, 65 years old and over). Then, *d*_*aig*_ is the number of prescriptions of antibiotic class *a* associated with diagnosis code *i* in age group *g*, 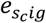 is the proportion of cases with diagnosis code *i* that are estimated to be caused by causative species *s*_*c*_ in age group *g*, and 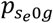 is the healthy carriage prevalence of exposed species *s*_*e*_ among individuals in age group *g*. When *s*_*c*_ and *s*_*e*_ are the same organism, 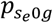 is set to 1. For each variable, we use the same data sources and estimation methods described in the main text (NAMCS/NHAMCS 2015, Human Microbiome Project, and published literature).

The diagonals, shaded in light gray, are “target” antibiotic exposures for the pathogen of interest; in other words, these are antibiotic exposures that the organism experienced because it was causing disease. The “Overall” column is all exposures to the antibiotic class of interest experienced by each species and is equivalent to *T*_*as*_ in Equation 1 of the main text. The “Other” column is the difference between the “Overall” column and the sum of the remaining columns. Note that this column represents exposures due to non-bacterial causes as well as bacterial organisms other than our nine potential pathogens of interest.

The “Any antibiotic” table includes all prescriptions for drugs classified as antibiotics based on the Lexicon Plus® Therapeutic Classification Scheme (*Lexicon Plus*, 2008). Any drug with a Level 1 description of “anti-infectives”, but with a Level 2 description that was not “amebicides”, “antihelmintics”, “antifungals”, “antimalarial agents”, or “antiviral agents” was included as an antibiotic. The “Any included antibiotic class” table includes all prescriptions for drugs in the classes shown in the following tables. Antibiotic classes were defined using the Level 2 categories in Lexicon Plus®.

These tables are designed to facilitate a better understanding of the “nuts and bolts” of this work and to enable calculation of other metrics of interest. For example, the proportion of bystander exposures as calculated by the authors in an earlier paper (Tedijanto et al., 2018) for any given antibiotic and species pair can be calculated as one minus the target exposures (diagonal value) over the “Overall” value. These tables can also be used to understand how interventions focused on a single species (e.g. vaccines) may reduce antibiotic exposures experienced by another species.

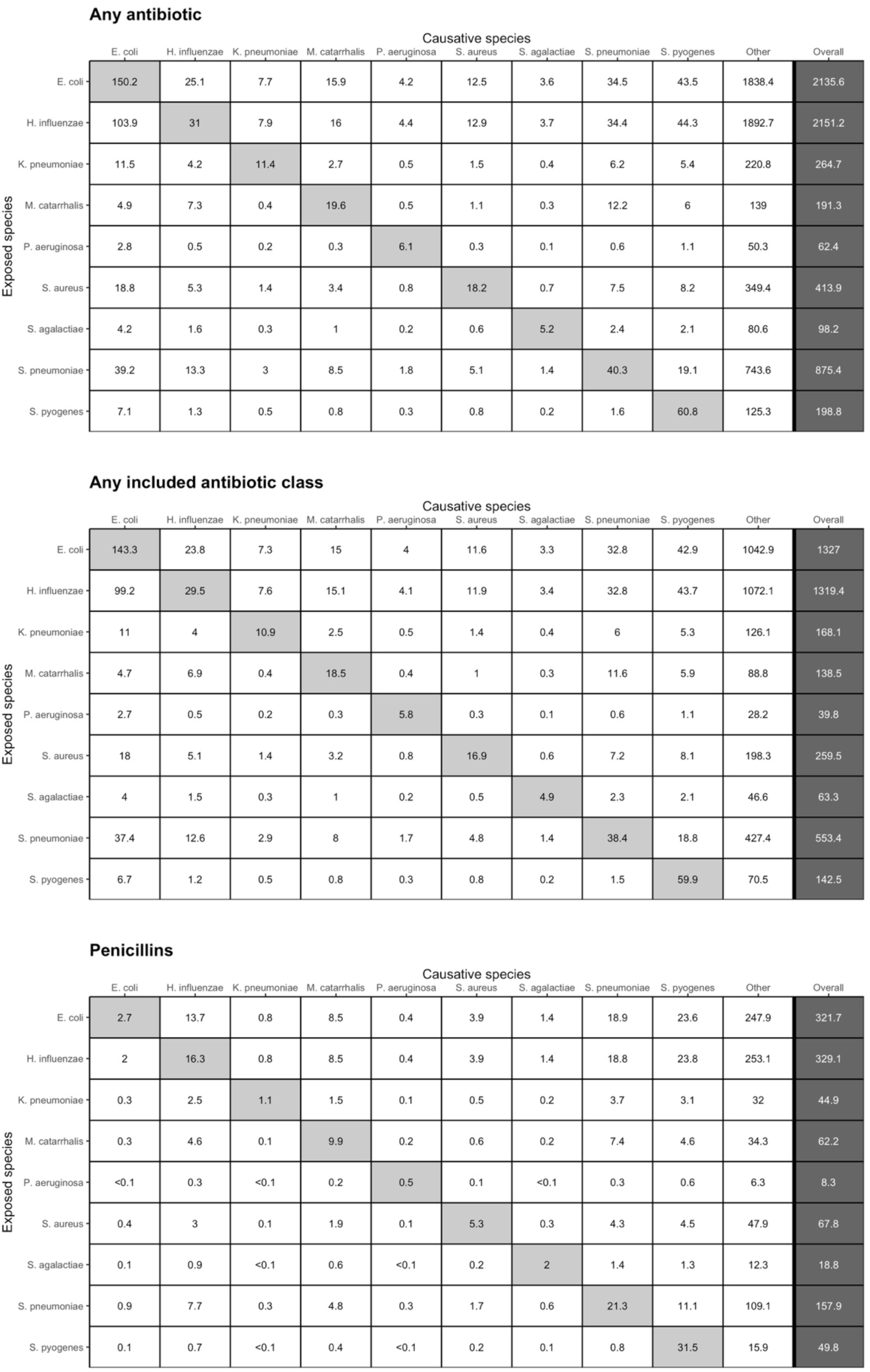
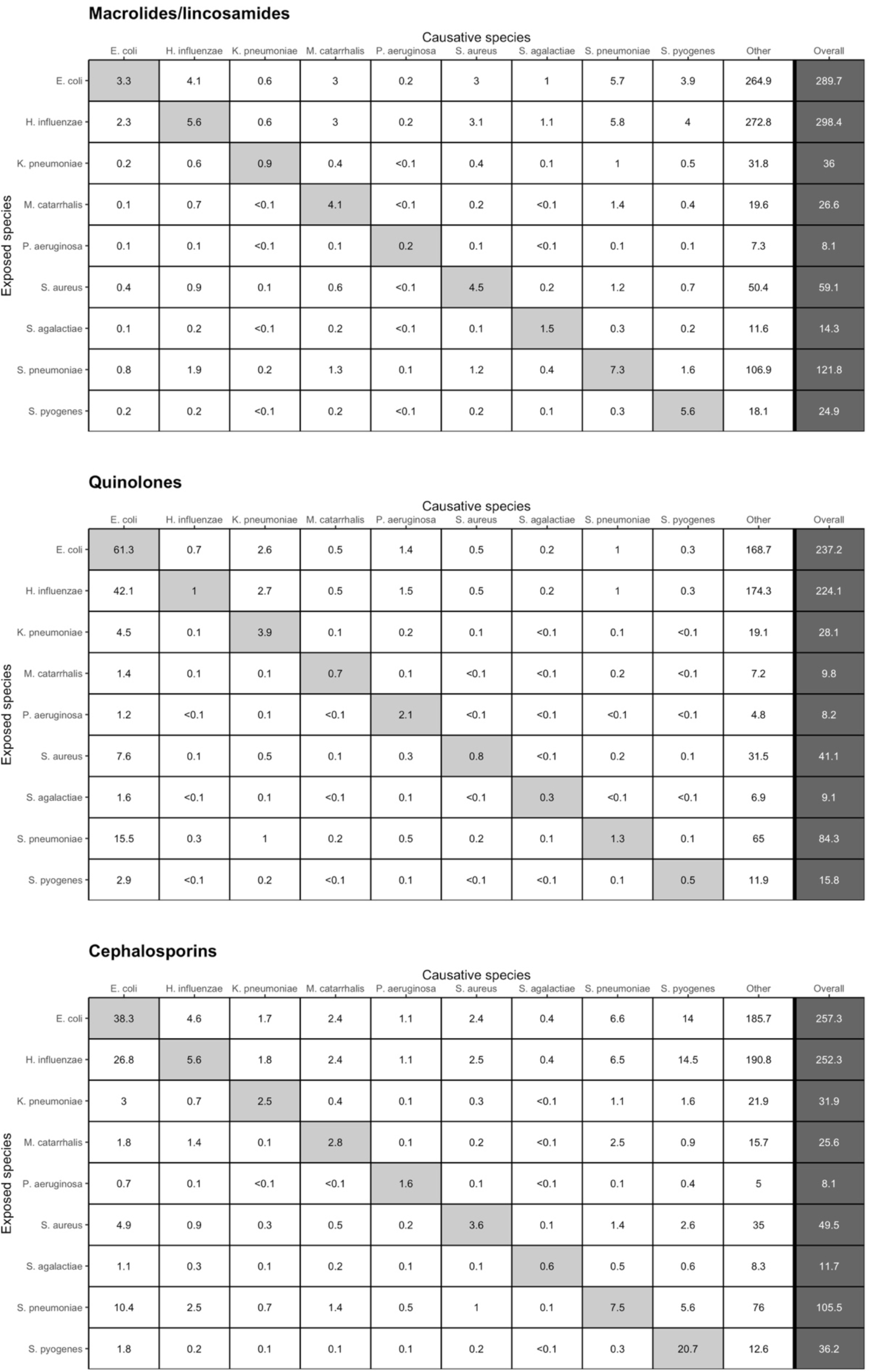
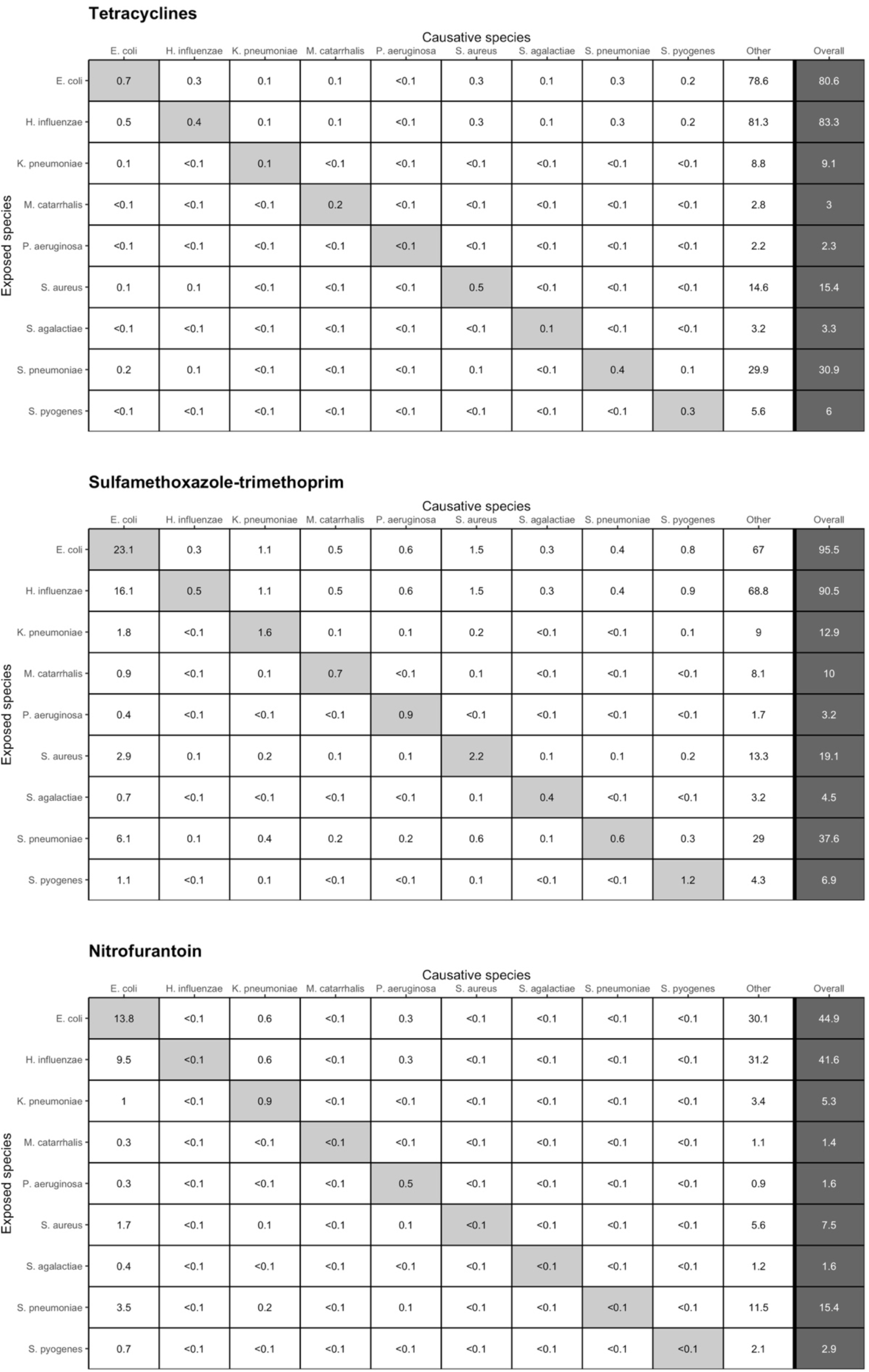

## Notes

### Author Declarations

All relevant ethical guidelines have been followed and any necessary IRB and/or ethics committee approvals have been obtained.

Any clinical trials involved have been registered with an ICMJE-approved registry such as ClinicalTrials.gov and the trial ID is included in the manuscript.

## References

Aitken, M., & Taylor, J. A. (1998). Prevalence of Clinical Sinusitis in Young Children Followed Up by Primary Care Pediatricians. Archives of Pediatrics & Adolescent Medicine, 152(3), 244–248. https://doi.org/10.1001/archpedi.152.3.244

Andersson, D. I., & Hughes, D. (2010). Antibiotic resistance and its cost: Is it possible to reverse resistance?Nature Reviews Microbiology, 8(4), 260–271. https://doi.org/10.1038/nrmicro2319

Bäckhed, F., Roswall, J., Peng, Y., Feng, Q., Jia, H., Kovatcheva-Datchary, P., … Wang, J. (2015). Dynamics and Stabilization of the Human Gut Microbiome during the First Year of Life. Cell Host & Microbe, 17(5), 690–703. https://doi.org/10.1016/j.chom.2015.04.004

Benninger, M. S., Holzer, S. E. S., & Lau, J. (2000). Diagnosis and treatment of uncomplicated acute bacterial rhinosinusitis: Summary of the Agency for Health Care Policy and Research evidence-based report. Otolaryngology–Head and Neck Surgery, 122(1), 1–7. https://doi.org/10.1016/S0194-5998(00)70135-5

Bluestone, C. D., Stephenson, J. S., & Martin, L. M. (1992). Ten-year review of otitis media pathogens. The Pediatric Infectious Disease Journal, 11(8), S7–S11. https://doi.org/10.1097/00006454-199208001-00002

Bogaert, D., Keijser, B., Huse, S., Rossen, J., Veenhoven, R., Gils, E. V., … Sanders, E. (2011). Variability and Diversity of Nasopharyngeal Microbiota in Children: A Metagenomic Analysis. PLoS ONE, 6(2), e17035. https://doi.org/10.1371/journal.pone.0017035

Brook, I. (2016). Microbiology of chronic rhinosinusitis. European Journal of Clinical Microbiology & Infectious Diseases, 35(7), 1059–1068. https://doi.org/10.1007/s10096-016-2640-x

Brook, I., Thompson, D. H., & Frazier, E. H. (1994). Microbiology and Management of Chronic Maxillary Sinusitis. Archives of Otolaryngology–Head & Neck Surgery, 120(12), 1317–1320.

Buffie, C. G., & Pamer, E. G. (2013). Microbiota-mediated colonization resistance against intestinal pathogens. Nature Reviews Immunology, 13(11), 790–801. https://doi.org/10.1038/nri3535

Burgstaller, J. M., Steurer, J., Holzmann, D., Geiges, G., & Soyka, M. B. (2016). Antibiotic efficacy in patients with a moderate probability of acute rhinosinusitis: A systematic review. European Archives of Oto-Rhino-Laryngology, 273(5), 1067–1077. https://doi.org/10.1007/s00405-015-3506-z

Celin, S. E., Bluestone, C. D., Stephenson, J., Yilmaz, H. M., & Collins, J. J. (1991). Bacteriology of Acute Otitis Media in Adults. JAMA, 266(16), 2249–2252. https://doi.org/10.1001/jama.266.16.2249

Chira, S., & Miller, L. G. (2010). Staphylococcus aureus is the most common identified cause of cellulitis: A systematic review. Epidemiology and Infection, 138(3), 313–317. https://doi.org/10.1017/S0950268809990483

Chua, K.-P., Fischer, M. A., & Linder, J. A. (2019). Appropriateness of outpatient antibiotic prescribing among privately insured US patients: ICD-10-CM based cross sectional study. BMJ, 364(k5092). https://doi.org/10.1136/bmj.k5092

Durkin, M. J., Jafarzadeh, S. R., Hsueh, K., Sallah, Y. H., Munshi, K. D., Henderson, R. R., & Fraser, V. J. (2018). Outpatient Antibiotic Prescription Trends in the United States: A National Cohort Study. Infection Control & Hospital Epidemiology, 39(5), 584–589. https://doi.org/10.1017/ice.2018.26

Edlin, R. S., Shapiro, D. J., Hersh, A. L., & Copp, H. L. (2013). Antibiotic resistance patterns of outpatient pediatric urinary tract infections. The Journal of Urology, 190(1), 222–227. https://doi.org/10.1016/j.juro.2013.01.069

European Centre for Disease Prevention and Control. (2018). Antimicrobial consumption: Annual epidemiological report for 2017. Stockholm.

Filice, G. A., Drekonja, D. M., Thurn, J. R., Hamann, G. M., Masoud, B. T., & Johnson, J. R. (2015). Diagnostic Errors that Lead to Inappropriate Antimicrobial Use. Infection Control & Hospital Epidemiology, 36(8), 949–956. https://doi.org/10.1017/ice.2015.113

Fleming-Dutra, K. E., Hersh, A. L., Shapiro, D. J., Bartoces, M., Enns, E. A., File, T. M., … Hicks, L. A. (2016). Prevalence of Inappropriate Antibiotic Prescriptions Among US Ambulatory Care Visits, 2010-2011. JAMA, 315(17), 1864–1873. https://doi.org/10.1001/jama.2016.4151

Fokkens, W., Lund, V., & Mullol, J. (2007). European position paper on rhinosinusitis and nasal polyps 2007. A summary for otorhinolaryngologists. Rhinology, 45(97).

Ginsburg, C. M., McCracken, G. H., Crow, S. D., Dildy, B. R., Morchower, G., Steinberg, J. B., & Lancaster, K. (1985). Seroepidemiology of the Group-A Streptococcal Carriage State in a Private Pediatric Practice. American Journal of Diseases of Children, 139(6), 614–617. https://doi.org/10.1001/archpedi.1985.02140080084039

Gunnarsson, R. K., Holm, S. E., & Söderström, M. (1997). The prevalence of beta-haemolytic streptococci in throat specimens from healthy children and adults: Implications for the clinical value of throat cultures. Scandinavian Journal of Primary Health Care, 15(3), 149–155. https://doi.org/10.3109/02813439709018506

Gupta, K., Hooton, T. M., Naber, K. G., Wullt, B., Colgan, R., Miller, L. G., … Soper, D. E. (2011). International Clinical Practice Guidelines for the Treatment of Acute Uncomplicated Cystitis and Pyelonephritis in Women: A 2010 Update by the Infectious Diseases Society of America and the European Society for Microbiology and Infectious Diseases. Clinical Infectious Diseases, 52(5), e103–e120. https://doi.org/10.1093/cid/ciq257

Gupta, K., Scholes, D., & Stamm, W. E. (1999). Increasing prevalence of antimicrobial resistance among uropathogens causing acute uncomplicated cystitis in women. JAMA, 281(8), 736–738. https://doi.org/10.1001/jama.281.8.736

Gwaltney, Jack M., Scheld, W. M., Sande, M. A., & Sydnor, A. (1992). The Microbial Etiology and Antimicrobial Therapy of Adults with Acute Community-Acquired Sinusitis—A 15-Year Experience at the University of Virginia and Review of Other Selected Studies. Journal of Allergy and Clinical Immunology, 90(3), 457–462. https://doi.org/10.1016/0091-6749(92)90169-3

Hammitt, L. L., Bruden, D. L., Butler, J. C., Baggett, H. C., Hurlburt, D. A., Reasonover, A., & Hennessy, T. W. (2006). Indirect Effect of Conjugate Vaccine on Adult Carriage of Streptococcus pneumoniae: An Explanation of Trends in Invasive Pneumococcal Disease. The Journal of Infectious Diseases, 193(11), 1487–1494. https://doi.org/10.1086/503805

Holgerson, P. L., Öhman, C., Rönnlund, A., & Johansson, I. (2015). Maturation of oral microbiota in children with or without dental caries. PLoS ONE, 10(5), 1–20. https://doi.org/10.1371/journal.pone.0128534

Hooton, T. M., & Gupta, K. (2019). Acute simple cystitis in women. In UpToDate, Post, TW (Ed). Retrieved from https://www.uptodate.com/contents/acute-simple-cystitis-in-women

Huang, Susan S., Hinrichsen, V. L., Stevenson, A. E., Rifas-Shiman, S. L., Kleinman, K., Pelton, S. I., … Finkelstein, J. A. (2009). Continued Impact of Pneumococcal Conjugate Vaccine on Carriage in Young Children. Pediatrics, 124(1), e1–e11. https://doi.org/10.1542/peds.2008-3099

Huttenhower, C., Gevers, D., Knight, R., Abubucker, S., Badger, J. H., Chinwalla, A. T., … White, O. (2012). Structure, function and diversity of the healthy human microbiome. Nature, 486(7402), 207–214. https://doi.org/10.1038/nature11234

Jain, S., Self, W. H., Wunderink, R. G., Fakhran, S., Balk, R., Bramley, A. M., … Finelli, L. (2015). Community-Acquired Pneumonia Requiring Hospitalization among U.S. Adults. New England Journal of Medicine, 373(5), 415–427. https://doi.org/10.1056/NEJMoa1500245

Kabbani, S., Hersh, A. L., Shapiro, D. J., Fleming-Dutra, K. E., Pavia, A. T., & Hicks, L. A. (2018). Opportunities to Improve Fluoroquinolone Prescribing in the United States for Adult Ambulatory Care Visits. Clinical Infectious Diseases, 67(1), 134–136. https://doi.org/10.1093/cid/ciy035

Khosravi, A., & Mazmanian, S. K. (2013). Disruption of the gut microbiome as a risk factor for microbial infections. Current Opinion in Microbiology, 16(2), 221–227. https://doi.org/10.1016/j.mib.2013.03.009

King, L. M., Bartoces, M., Fleming-Dutra, K. E., Roberts, R. M., & Hicks, L. A. (2019). Changes in US Outpatient Antibiotic Prescriptions from 2011-2016. Clinical Infectious Diseases. https://doi.org/10.1093/cid/ciz225

Klevens, R. M., Caten, E., Olesen, S. W., DeMaria, A., Troppy, S., & Grad, Y. H. (2019). Outpatient antibiotic prescribing in Massachusetts, 2011-2015. Open Forum Infectious Diseases, 6(5), ofz169. https://doi.org/10.1093/ofid/ofz169

Lexicon Plus. (2008). Retrieved from https://www.cerner.com/solutions/drug-database

Linder, J. A. (2008). Editorial Commentary: Antibiotics for Treatment of Acute Respiratory Tract Infections: Decreasing Benefit, Increasing Risk, and the Irrelevance of Antimicrobial Resistance. Clinical Infectious Diseases, 47(6), 744–746. https://doi.org/10.1086/591149

Lipsitch, M. (2001). The rise and fall of antimicrobial resistance. Trends in Microbiology, 9(9), 438–444. https://doi.org/10.1016/S0966-842X(01)02130-8

MacLean, R. C., & San Millan, A. (2019). The evolution of antibiotic resistance. Science, 365(6458), 1082–1083. https://doi.org/10.1126/science.aax3879

Mainous, Arch G., Hueston, W. J., Everett, C. J., & Diaz, V. A. (2006). Nasal Carriage of Staphylococcus aureus and Methicillin-Resistant S aureus in the United States, 2001-2002. The Annals of Family Medicine, 4(2), 132–137. https://doi.org/10.1370/afm.526

National Center for Health Statistics. (2015a). National Ambulatory Medical Care Survey—Public-use data file and documentation. Retrieved from ftp://ftp.cdc.gov/pub/Health_Statistics/NCHS/Datasets/NAMCS

National Center for Health Statistics. (2015b). National Hospital Ambulatory Medical Care Survey—Public-use data file and documentation. Retrieved from ftp://ftp.cdc.gov/pub/Health_Statistics/NCHS/Datasets/NHAMCS

Olesen, S. W., Barnett, M. L., MacFadden, D. R., Brownstein, J. S., Hernández-Díaz, S., Lipsitch, M., & Grad, Y. H. (2018). The distribution of antibiotic use and its association with antibiotic resistance. ELife, 7(e39435). https://doi.org/10.7554/eLife.39435.001

Olesen, S. W., Barnett, M. L., MacFadden, D. R., Lipsitch, M., & Grad, Y. H. (2018). Trends in outpatient antibiotic prescribing practice among US older adults, 2011-2015: An observational study. BMJ, 362, k3155. https://doi.org/10.1136/bmj.k3155

Pettigrew, M. M., Laufer, A. S., Gent, J. F., Kong, Y., Fennie, K. P., & Metlay, J. P. (2012). Upper respiratory tract microbial communities, acute otitis media pathogens, and antibiotic use in healthy and sick children. Applied and Environmental Microbiology, 78(17), 6262–6270. https://doi.org/10.1128/AEM.01051-12

Pouwels, K. B., Batra, R., Patel, A., Edgeworth, J. D., Robotham, J. V., & Smieszek, T. (2017). Will co-trimoxazole resistance rates ever go down? Resistance rates remain high despite decades of reduced co-trimoxazole consumption. Journal of Global Antimicrobial Resistance, 11, 71–74. https://doi.org/10.1016/j.jgar.2017.07.013

Public Health Agency of Canada. (2018). Canadian Antimicrobial Resistance Surveillance System 2017 Report.

Public Health England. (2017). English Surveillance Programme for Antimicrobial Utilisation and Resistance (ESPAUR) 2017.

Regev-Yochay, G., Dagan, R., Raz, M., Carmeli, Y., Shainberg, B., Derazne, E., … Rubinstein, E. (2004). Association Between Carriage of Streptococcus pneumoniae and Staphylococcus aureus in Children. JAMA, 292(6), 716–720. https://doi.org/10.1001/jama.292.6.716

Rosenfeld, R. M., Piccirillo, J. F., Chandrasekhar, S. S., Brook, I., Kumar, K. A., Kramper, M., … Corrigan, M. D. (2015). Clinical Practice Guideline (Update): Adult Sinusitis. Otolaryngology–Head and Neck Surgery, 152(2S), S1–S39. https://doi.org/10.1177/0194599815572097

Sakoulas, G., Geriak, M., & Nizet, V. (2018). Is a Reported Penicillin Allergy Sufficient Grounds to Forgo the Multidimensional Antimicrobial Benefits of β-Lactam Antibiotics?Clinical Infectious Diseases, 68(1), 157–164. https://doi.org/10.1093/cid/ciy557

Sande, M. A., & Gwaltney, J. M. (2004). Acute Community-Acquired Bacterial Sinusitis: Continuing Challenges and Current Management. Clinical Infectious Diseases, 39, S151–S158. https://doi.org/10.1086/421353

Shehab, N., Patel, P. R., Srinivasan, A., & Budnitz, D. S. (2008). Emergency Department Visits for Antibiotic-Associated Adverse Events. Clinical Infectious Diseases, 47(6), 735–743. https://doi.org/10.1086/591126

Smith, S. S., Evans, C. T., Tan, B. K., Chandra, R. K., Smith, S. B., & Kern, R. C. (2013). National burden of antibiotic use for adult rhinosinusitis. Journal of Allergy and Clinical Immunology, 132(5), 1230–1232. https://doi.org/10.1016/j.jaci.2013.07.009

Snow, V., Mottur-Pilson, C., & Hickner, J. M. (2001). Principles of Appropriate Antibiotic Use for Acute Sinusitis in Adults. Annals of Internal Medicine, 134(6), 495. https://doi.org/10.7326/0003-4819-134-6-200103200-00016

Stewardson, A. J., Gaïa, N., François, P., Malhotra-Kumar, S., Delémont, C., Martinez de Tejada, B., … Lazarevic, V. (2015). Collateral damage from oral ciprofloxacin versus nitrofurantoin in outpatients with urinary tract infections: A culture-free analysis of gut microbiota. Clinical Microbiology and Infection, 21(4), 344.e1-344.e11. https://doi.org/10.1016/j.cmi.2014.11.016

Swedres-Svarm. (2017). Consumption of antibiotics and occurrence of antibiotic resistance in Sweden. Public Health Agency of Sweden and National Veterinary Institute.

Tedijanto, C., Olesen, S. W., Grad, Y. H., & Lipsitch, M. (2018). Estimating the proportion of bystander selection for antibiotic resistance among potentially pathogenic bacterial flora. Proceedings of the National Academy of Sciences, 115(51), E11988–E11995. https://doi.org/10.1073/pnas.1810840115

The Human Microbiome Project Consortium. (2012). A framework for human microbiome research. Nature, 486(7402), 215–221. https://doi.org/10.1038/nature11209

Tobia, C. C., Aspinall, S. L., Good, C. B., Fine, M. J., & Hanlon, J. T. (2008). Appropriateness of antibiotic prescribing in veterans with community-acquired pneumonia, sinusitis, or acute exacerbations of chronic bronchitis: A cross-sectional study. Clinical Therapeutics, 30(6), 1135–1144. https://doi.org/10.1016/j.clinthera.2008.06.009

Tomas, M. E., Getman, D., Donskey, C. J., & Hecker, M. T. (2015). Overdiagnosis of Urinary Tract Infection and Underdiagnosis of Sexually Transmitted Infection in Adult Women Presenting to an Emergency Department. Journal of Clinical Microbiology, 53(8), 2686–2692. https://doi.org/10.1128/JCM.00670-15

Vanden Eng, J., Marcus, R., Hadler, J. L., Imhoff, B., Vugia, D. J., Cieslak, P. R., … Besser, R. E. (2003). Consumer Attitudes and Use of Antibiotics. Emerging Infectious Diseases, 9(9), 1128–1135. https://doi.org/10.3201/eid0909.020591

Verhaegh, S. J. C., Lebon, A., Saarloos, J. A., Verbrugh, H. A., Jaddoe, V. W. V., Hofman, A., … van Belkum, A. (2010). Determinants of Moraxella catarrhalis colonization in healthy Dutch children during the first 14 months of life. Clinical Microbiology and Infection, 16(7), 992–997. https://doi.org/10.1111/j.1469-0691.2009.03008.x

Wald, E. R., Guerra, N., & Byers, C. (1991). Upper Respiratory Tract Infections in Young Children: Duration of and Frequency of Complications. Pediatrics, 87(2), 129–133.

Williams, J. W., Simel, D. L., Roberts, L., & Samsa, G. P. (1992). Clinical Evaluation for Sinusitis. Annals of Internal Medicine, 117(9), 705–710. https://doi.org/10.7326/0003-4819-117-9-705

Wubbel, Loretta, Muniz, L., Ahmed, A., Trujillo, M., Carubelli, C., McCoig, C., … McCracken, G. H. (1999). Etiology and treatment of community-acquired pneumonia in ambulatory children. The Pediatric Infectious Disease Journal, 18(2), 98–104.

Yassour, M., Vatanen, T., Siljander, H., Hämäläinen, A.-M., Härkönen, T., Ryhänen, S. J., … Xavier, R. J. (2016). Natural history of the infant gut microbiome and impact of antibiotic treatments on strain-level diversity and stability. Science Translational Medicine, 8(343), 129–139. https://doi.org/10.1126/scitranslmed.aad0917

